# Differential Recovery Trajectories of Emergency Otolaryngologic Conditions across the COVID-19 Pandemic: A Six-year Longitudinal Study from an Urban Emergency Center

**DOI:** 10.64898/2026.06.20.26356151

**Authors:** Makoto Ogawa

**Author notes:** **Corresponding author:** Makoto Ogawa, Department of Otorhinolaryngology, Osaka Dental University, Otemae 1-5-17, Chuou-ku, Osaka, 540-0008, Japan.

## Abstract

**Objective:** The COVID-19 pandemic markedly altered social activity patterns, healthcare utilization, and the epidemiology of infectious diseases. However, its long-term impact on emergency otolaryngologic conditions remains incompletely understood. This study investigated long-term trends in emergency otolaryngologic conditions before, during, and after the COVID-19 pandemic using comprehensive data from a large urban emergency clinic in Osaka, Japan.

**Methods:** All new otolaryngologic outpatients who visited the Chuo Emergency Medical Clinic (CEMC) in Osaka City between 2019 and 2024were retrospectively analyzed. Annual trends in absolute numbers and relative proportions of emergency otolaryngologic conditions were examined by anatomical region and disease category, using 2019 as the pre-pandemic baseline.

**Results:** A total of 99,324 new otolaryngologic outpatients were analyzed. Overall emergency visits declined sharply to approximately half of baseline in 2020, followed by a gradual but incomplete recovery toward pre-pandemic levels by 2024. Most anatomical categories declined to 45–61% of baseline in 2020 and exhibited gradual yet incomplete recovery through 2023; in stark contrast, laryngeal conditions diverged sharply, surging beyond pre-pandemic levels after 2022. Acute infectious otorhinolaryngologic diseases fell to 23–50% of baseline in 2020 and showed variable recovery (69–103%) by 2024. Notably, laryngitis exceeded the baseline, reaching 132% in 2023, whereas epiglottic edema exhibited only a transient increase approaching the baseline in 2021. Non-infectious emergency conditions generally showed only a marginal decrease in 2020 and remained relatively stable throughout the study period, except for sudden sensorineural hearing loss (SSNHL), which dropped sharply to 39% of the baseline in 2020 and remained persistently reduced through 2024. Traumatic emergencies declined variably to 53-81% of the baseline in 2020, followed by an incomplete recovery, reaching only 55-69% by 2024.

**Conclusion:** Emergency otolaryngologic conditions demonstrated heterogeneous recovery trajectories following the COVID-19 pandemic. While most infectious and traumatic conditions gradually but incompletely normalized, laryngeal conditions showed a distinct post-pandemic surge, and SSNHL remained persistently suppressed. These findings reveal heterogeneous, condition-specific recovery trajectories that reflect both genuine shifts in community pathogen burden, true traumatic incidence, and persistent alterations in healthcare-seeking behaviors—insights essential for resource allocation during future public health emergencies.

## 1. Introduction

The COVID-19 pandemic profoundly altered healthcare utilization worldwide, driven by large-scale public health restrictions, behavioral modifications, and the reduced circulation of common infectious pathogens. Notably, in the field of otolaryngology, marked reductions in outpatient volumes and elective surgical interventions were reported immediately following the onset of the pandemic [1–7], accompanied by substantial declines in acute upper respiratory tract infections [8–13].

However, while the previous hospital-based studies [1–13] provide valuable insights into elective care trajectories and the numbers of patients with various otolaryngologic conditions, most remain limited to the immediate onset or early phase of the pandemic. In contrast, our previous study [7] stands out as the only longitudinal investigation to have tracked the clinical trajectories of both surgical and referral patients with identical disease entities through a comprehensive timeline encompassing the pre-, mid-, and post-pandemic phases up to the eventual convergence. Specifically, our previous study [7] demonstrated that pediatric elective surgeries, such as adenoidectomy and tonsillectomy, as well as the referrals remained suppressed immediately after the pandemic’s onset and throughout the mid-pandemic period, but experienced an unprecedented, abrupt surge immediately following the relaxation of public health restrictions and the governmental reclassification of COVID-19 in Japan. This explosive post-restriction rebound in surgical and referral cases strongly suggests that young children and their caregivers who had hesitated to access medical institutions to avoid COVID-19 infection inherently desired to undergo these surgeries; furthermore, it implies that the temporary suppression and subsequent sudden resurgence in respiratory pathogens following the implementation and eventual cancellation of public health interventions triggered a synchronized wave of “catch-up” adenotonsillar hypertrophy, particularly in a cohort of young children who had spent their critical immunological development periods isolated from common community infections. However, hospital-based surgical volumes inherently conflate two distinct signals: shifts in healthcare-seeking behavior and genuine fluctuations in disease incidence. To accurately analyze these factors, it may be effective to use data from an emergency setting where patients present with intolerable or urgent symptoms. Such an approach minimizes the bias of patients choosing to avoid hospital visits.

Emergency otolaryngologic care represents a unique clinical setting in which patients present with a broad spectrum of acute conditions requiring prompt medical attention, including severe infections, epistaxis, trauma, and airway-threatening foreign bodies. During the early phase of the COVID-19 pandemic, elective clinical services and routine outpatient visits declined markedly [1–7], since patients with mild or manageable symptoms avoided seeking medical care and non-urgent procedures were postponed. In contrast, emergency otolaryngologic conditions are typically associated with intolerable or urgent symptoms that necessitate immediate medical intervention. It has therefore been hypothesized that patient volumes at specialized emergency medical centers would be less susceptible to pandemic-related behavioral changes than those at routine outpatient facilities or acute care hospitals.

Nevertheless, several studies have reported that the overall volume of emergency department visits and ambulance requests decreased during the early phase of the pandemic [14–18], accompanied by both quantitative and qualitative shifts in emergency care [16,18]. For instance, Katayama et al. [18] reported that in Osaka Prefecture, Japan, the number of ambulance requests decreased in 2020 and 2021, whereas the severity and mortality of hospitalized emergency patients increased during the same period. In the field of otolaryngology, several investigators have similarly noted a marked decrease in emergency department presentations during the early phase of the pandemic [19–22]. However, none of these existing emergency-specific studies have tracked the full multi-year longitudinal trajectories in case volumes spanning the pre-, mid-, and post-pandemic phases up to the eventual convergence.

To overcome these limitations, a specialized clinical setting that systematically consolidates regional urgent care is required. In Japan, emergency otolaryngologic care systems vary markedly across regions. In many prefectures, patients who develop emergency otolaryngologic symptoms during nighttime hours or holidays are evaluated by on-duty otolaryngologists at general hospitals on a rotating basis. This decentralized structure often leads to substantial physician fatigue and significant regional disparities in emergency otolaryngological coverage.

In contrast, the Chuo Emergency Medical Clinic (CEMC) is operated under the auspices of Osaka City, where otolaryngologic services are continuously staffed by rotating board-certified specialists. This coordinated system mitigates excessive burdens on individual clinicians and ensures that patients across the metropolitan area with severe or intolerable symptoms are consistently directed to a single emergency-designated center. Consequently, the CEMC provides an uncommonly comprehensive, population-level dataset that directly reflects the community-level burden, genuine disease incidence, and healthcare-seeking behaviors for emergent ENT diseases—a clinical resource that is rarely available in other regions of Japan or internationally.

Leveraging this unique, multi-year dataset from the CEMC, the present study aimed to examine longitudinal changes in emergency otolaryngological disease patterns, such as infectious diseases, representative functional disorders and traumatic injury, encompassing the pre-, mid-, and post-pandemic phases. Specifically, by analyzing both absolute numbers and relative proportions of emergency visits across anatomical regions and specific disease categories over a six-year period from 2019 to 2024, this study sought to delineate condition-specific recovery trajectories and to determine whether emergency infectious otolaryngologic diseases exhibited a post-pandemic “catch-up” surge analogous to that observed in elective adenotonsillar surgery.

## 2. Methods

### 2.1. Study Design and Setting

This retrospective observational study utilized data from the CEMC, which comprises four clinical departments: internal medicine, pediatrics, ophthalmology, and otolaryngology. As a central primary urgent facility, it manages a high annual workload consisting of both walk-in outpatients with acute conditions and emergency ambulance transports. The majority of patients presenting to the CEMC reside within Osaka City or its neighboring municipalities, such as Suita Toyonaka, Higashi-osaka, Yao, and Sakai, although a small proportion travel from more distant areas within Osaka Prefecture, or adjacent prefectures. To contextualize the scale of this regional cohort, as of 2025, the estimated population of Osaka City was approximately 2.82 million, while this practical commuter-based catchment area encompassing the contiguous neighboring cities represents a substantial population of over 5 million.

Otolaryngologic care at the CEMC is provided by rotating board-certified specialists from regional hospitals or private practices, ensuring consistent same-day evaluation and immediate management of acute ENT complaints. All patient encounters are systematically documented in a standardized electronic medical record system, which ensures the reliability of year-to-year comparisons of diagnostic trends. Furthermore, the aggregated annual clinical data are routinely published in the *Bulletin* o*f the Osaka Otolaryngologists’ Association*, serving as formalized external records that validate the continuity of diagnostic distributions across consecutive years.

The specific diagnostic categories utilized in this study, stratified by anatomical region, are comprehensively summarized in **Table 1**. To maintain diagnostic consistency and avoid overlapping counts, a strict data entry protocol is enforced at the center: for each emergency encounter, attending otolaryngologists are mandated to select only a single primary diagnosis. Although the record format allows for the documentation of secondary comorbidities, this primary code corresponds directly to the acute condition that most heavily contributed to the patient’s chief complaint or initial clinical presentation.

**Table 1.** Diagnostic categories stratified by anatomical region used in the Chuo Emergency Medical Center (CEMC). For each visit, otolaryngologists at CEMC recorded a single primary diagnosis corresponding to the condition most strongly contributing to the patient’s presenting complaint.

| Region |  |  |  |  |  |
| --- | --- | --- | --- | --- | --- |
| Ear | Nose | Oral cavity | Pharynx | Larynx | Trachea/Esophagus |
| Acute otitis media/mastoiditis | Epistaxis | Stomatitis | Pharyngitis/tonsillitis | Laryngitis | Foreign body (trachea) |
| Otitis externa | Acute rhinosinusitis | Abscess in the floor of the mouth | Peritonsillar abscess | Epiglottic edema | Foreign body (esophagus) |
| Catarrhal otitis media | Chronic rhinosinusitis | Salivary gland diseases | Postpharyngeal abscess | Injury | Others |
| Injury (ear canal) | Maxillary cyst | Oral injury | Foreign body | Foreign body |  |
| Injury (ear drum) | Nasal bone fracture | Neoplasms | Injury | Neoplasms |  |
| Foreign body (ear canal) | Foreign bodies (nasal cavity) | dental diseases | Neoplasms | Others |  |
| Vertigo/dizziness | Neoplasms | Others | Others |  |  |
| Sudden hearing loss | Others |  |  |  |  |
| Others |  |  |  |  |  |

The study period spanned from 2019 to 2024, covering the pre-, mid-, and post-pandemic phases of the COVID-19 era. The post-pandemic phase was operationally defined based on the governmental reclassification of COVID-19 to a Category V infectious disease under the Japanese Infectious Disease Control Law, which was announced in January 2023 and formally implemented in May 2023. The reclassification placed COVID-19 among routinely monitored infectious diseases, such as seasonal influenza, and marked a definitive transition toward the relaxation of public health restrictions and the normalization of clinical practices.

### 2.2. Data Acquisition and Subclassification Analysis

Annual longitudinal volumes of emergency otolaryngologic outpatient visits at the CEMC were evaluated using aggregated data published in the *Bulletin of the Osaka Otolaryngologists’ Association*. For the trend analysis, the corresponding electronic data were provided by the executive board members of the association.

For the subclassification analysis, outpatient visits were stratified based on the predefined diagnostic categories listed in **Table 1**, focusing on specific anatomical regions and distinct disease entities, including infectious diseases, traumatic injuries, and other representative otolaryngologic emergencies. Annual trends in relative proportions were subsequently evaluated for each category and individual disease, with the pre-pandemic year of 2019 established as the baseline value (2019 = 100).

### 2.3. Statistical Analysis

Given the comprehensive, population-based nature of this regional dataset and the study’s primary objective of describing long-term epidemiological trends, no formal inferential statistical hypothesis testing or *p*-value calculations were performed. Instead, longitudinal trajectories were assessed descriptively using absolute patient numbers and annual relative proportions, with the pre-pandemic year of 2019 as the baseline value.

## 3. Results

### 3.1 Overall Patients Volumes and Anatomical Distributions

A total of 99,324 new outpatients who visited the otolaryngology department at the CEMC during the six-year study period were included in the longitudinal analysis. As illustrated in **Fig. 1**, the annual volume of otolaryngologic visits declined sharply in 2020, coinciding with the onset of the COVID-19 pandemic. This immediate initial drop was followed by a gradual but incomplete recovery from 2021 through 2023 and a subsequent slight decrease in 2024. Throughout the entire study duration, conditions involving the otologic, rhinologic, and pharyngeal regions consistently comprised the vast majority of emergency encounters. In contrast, laryngeal diseases accounted for a substantially smaller proportion of the total clinic workload, while emergencies involving the oral cavity and trachea/esophagus consistently represented only minor fractions of the overall patient volume.

**Fig. 1.**
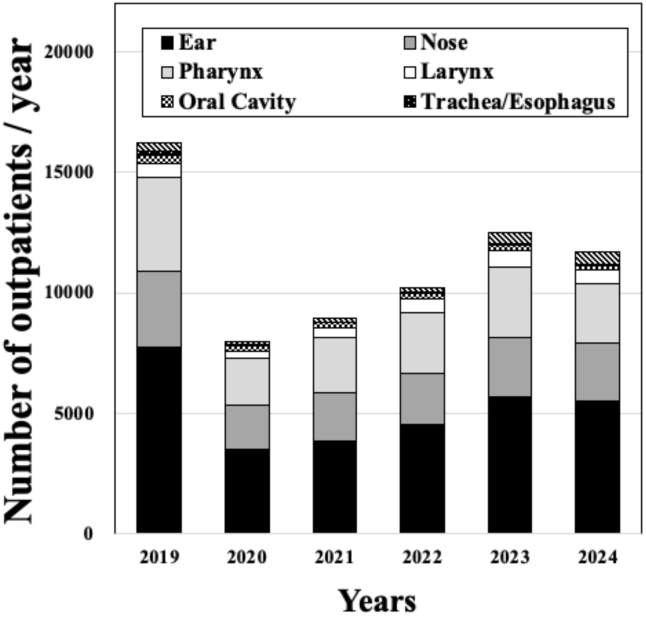
Annual trends in the number of new otolaryngologic outpatients at the Chuo Emergency Medical Center by anatomical region. The total number of visits decreased markedly in 2020, followed by a gradual recovery in subsequent years, approaching pre-pandemic levels by 2023 followed by a slight decrease in 2024.

### 3.2 Comparative Analysis of Anatomical Proportions

**Fig. 2** illustrates the longitudinal trends in the relative proportions of new otolaryngologic outpatient encounters at the CEMC stratified by anatomical region, using the pre-pandemic year of 2019 as the reference baseline (2019 = 100). In 2020, at the onset of the pandemic, all anatomical categories—with the notable exception of the larynx—exhibited a sharp decrease, dropping to 45%-61% of their baseline levels. This initial suppression was followed by a gradual but incomplete year-over-year recovery to approximately 75% of their baseline levels in 2023. In stark contrast, laryngeal conditions demonstrated a distinct and prominent upward trajectory beginning in 2022, surpassing the pre-pandemic baseline level in 2023.

**Fig. 2.**
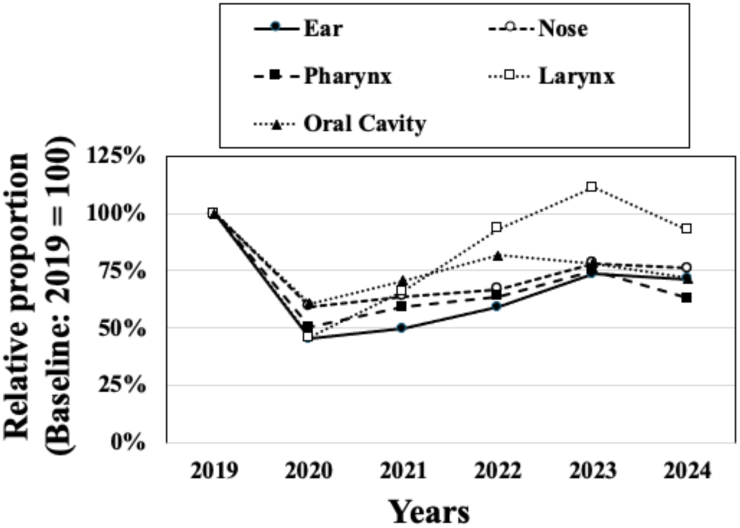
Annual trends in the relative proportions of new otolaryngologic outpatients at the Chuo Emergency Medical Center by anatomical region, using 2019 as the reference baseline (2019 = 100). In 2020, at the onset of the pandemic, all anatomical categories—with the notable exception of the larynx—exhibited a sharp decrease, followed by a gradual but incomplete increase in subsequent years, whereas laryngeal conditions uniquely demonstrated a sustained surge beginning in 2022.

### 3.3 Trends in Acute Infectious Diseases

**Fig. 3A** and **Fig. 3B** illustrate the annual trends in the absolute numbers and relative proportions of acute infectious otorhinolaryngologic diseases, respectively, using the pre-pandemic year of 2019 as the reference baseline (2019 = 100). At the pandemic’s onset (2020), all infectious conditions declined sharply, reaching only 23–50% of pre-pandemic levels (**Fig. 3B)**. Following the relaxation of public health restrictions, most disease categories — with the exception of laryngitis and epiglottic edema gradually increased, reaching 67%–81% of baseline levels by 2023. Individually, acute otitis media (AOM), acute sinusitis, and pharyngitis and tonsillitis recovered to 81%, 67%, and 68% of their baseline values. In stark contrast, laryngitis strikingly exceeded the baseline from 2022 onward. Meanwhile, epiglottic edema demonstrated a unique epidemiological pattern, characterized by a transient increase exclusively in 2021, followed by a gradual return to the levels of other infectious diseases except for laryngitis.

**Fig. 3.**
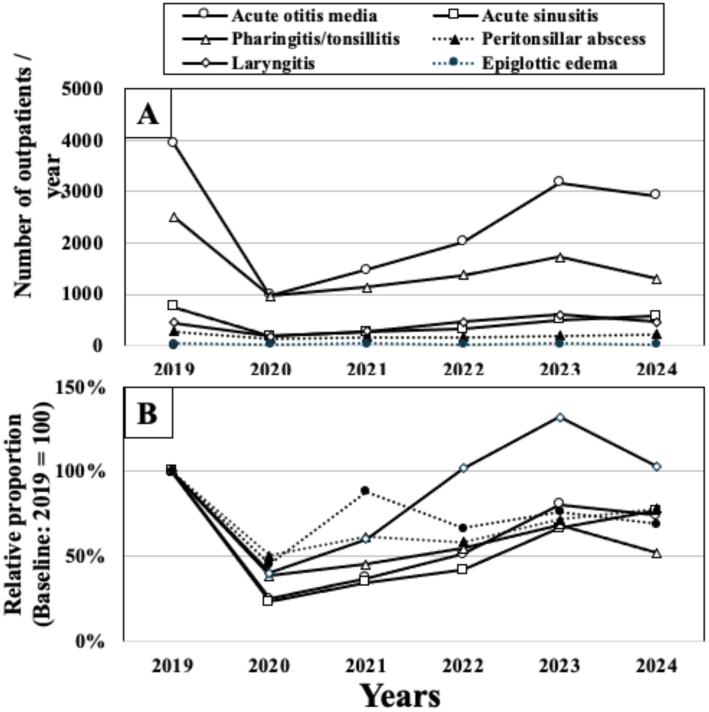
Annual trends in the absolute numbers and relative proportions of new outpatients with infectious otorhinolaryngologic diseases: (A) the absolute numbers, and (B) relative proportions using 2019 as the reference baseline (2019 = 100). Infectious diseases declined sharply in 2020, and all categories except laryngitis and epiglottic edema gradually increased after the relaxation of public health restrictions. In contrast, laryngitis exceeded the baseline from 2022 onward, whereas epiglottic edema exhibited a transient increase in 2021.

### 3.4 Trends in Representative Otolaryngologic Emergency Conditions

**Fig. 4A** and **Fig. 4B** show the annual trends in the absolute numbers and relative proportions of new outpatients presenting with representative otolaryngologic emergency conditions except trauma, using the pre-pandemic year of 2019 as the reference baseline (2019 = 100). In contrast to acute infectious diseases, all emergency conditions—with the exception of sensorineural sudden hearing loss (SSNHL) — exhibited only a marginal decrease in 2020 and remained relatively stable throughout the study period, showing modest year-to-year fluctuations. Notably, epistaxis and pharyngeal foreign body almost completely returned to their baseline levels by 2023, whereas vertigo and foreign bodies in the external ear and nasal cavity showed incomplete recovery, reaching only 57%-72% of their pre-pandemic baseline levels by 2024. However, only foreign bodies in the nasal cavity exhibited a gradual, continuous decline across the study period. Meanwhile, SSNHL exhibited the clearest and most continuous decline. The volume sharply dropped to 39% of the baseline in 2020 and remained at approximately 40% through 2024, showing a trajectory that clearly differed from other non-infectious emergencies.

**Fig. 4.**
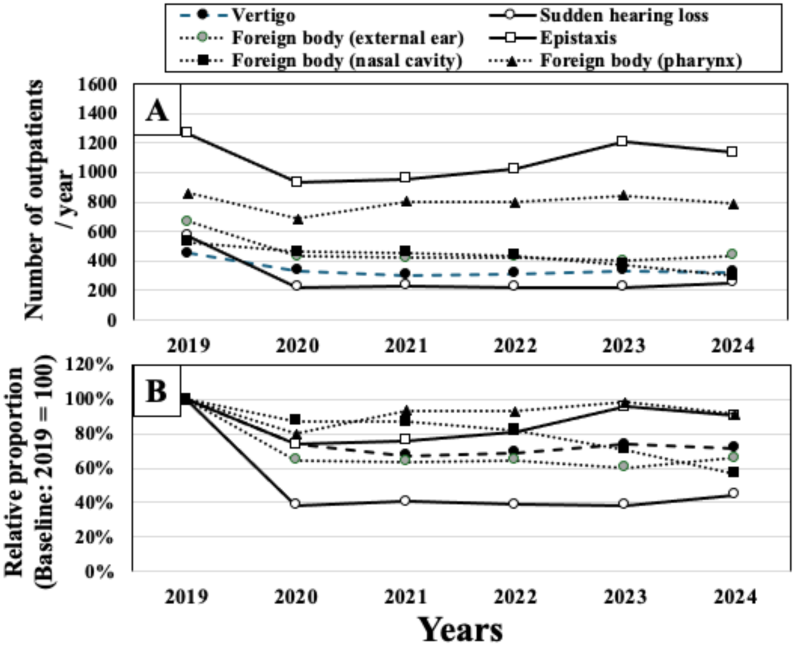
Annual trends in the absolute numbers and relative proponions of new outpatients with representative otorhinolaryngologic emergency conditions: (A) the absolute numbers, and (8) relative proponions using 2019 as the reference baseline (2019 = JOO). Emergency conditions, except sudden sensorineural hearing loss (SSNHL), showed relatively stable annual numbers throughout the study period, with only modest fluctuations compared with infectious diseases. By contrast, SSNHL exhibited a sharp decline in 2020 and subsequently remained persistently reduced.

### 3.5 Trends in Traumatic Emergency Conditions

**Fig. 5A** and **Fig. 5B** illustrate the annual trends in the absolute numbers and relative proportions of new outpatients presenting with traumatic emergency conditions by otorhinolaryngologic region, using the pre-pandemic year of 2019 as the reference baseline (2019 = 100). Although all categories declined in 2020, the magnitude of the decrease substantially varied by anatomical site, ranging from 53% to 81% of the baseline, followed by a gradual but incomplete recovery with variable fluctuations in subsequent years, reaching only 55-69% by 2024.

**Fig. 5.**
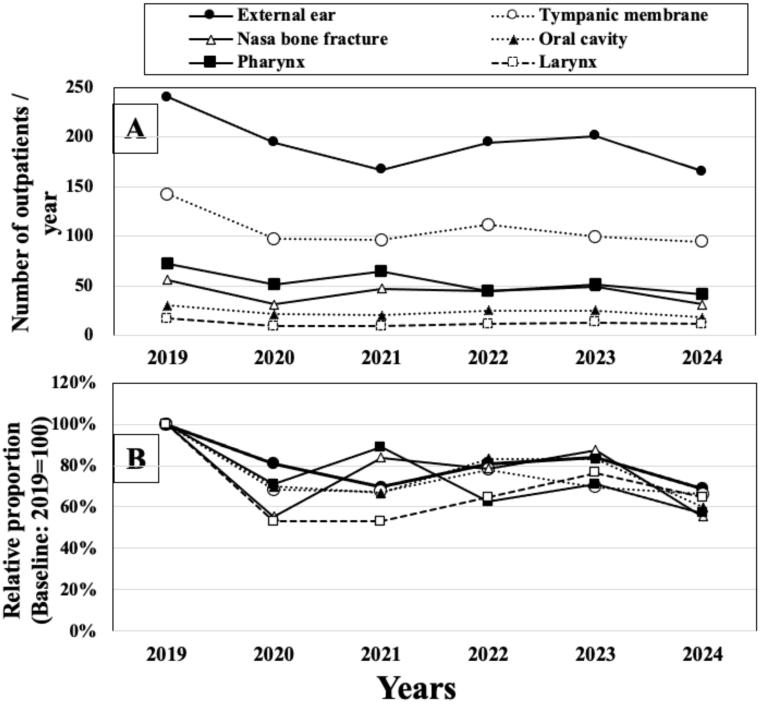
Annual trends in the numbers and relative proportions of new outpatients presenting with traumatic emergency conditions by otorhinolaryngologic region. (A) the absolute numbers, and (B) relative proportions using 2019 as the reference baseline (2019 = 100). In 2020, at the onset of the pandemic, all categories declined, whereas the magnitude of the decrease varied substantially by anatomical site, followed by a gradual but incomplete recovery with variable fluctuations in subsequent years.

## 4. Discussion

This six-year, single-center longitudinal study based on the dataset obtained from the CEMC, as a large urban emergency medical center, provides a comprehensive assessment of intra-pandemic declines and post-pandemic recovery patterns in the patient volumes of acute otorhinolaryngologic infections as well as representative and traumatic otolaryngologic emergency conditions. Crucially, because the CEMC specializes strictly in emergency care for patients who require immediate medical interventions during nighttime and holidays, the patient volumes are substantially less susceptible to pandemic-related behavioral changes— such as voluntary medical avoidance or shifts in elective care-seeking behaviors—than routine outpatient clinics. Consequently, the longitudinal data presented here uniquely reflect the genuine community-level pathogen burden, genuine infectious incidence, true traumatic incidence, and broader epidemiological and social dynamics during the health crisis with minimal confounding noise.

### 4.1 An Immediate Sharp Volume Decline and Subsequent Different Recovery by Anatomical Region

Consistent with numerous previous reports [1–22], the total patient volumes for emergency otolaryngologic conditions at the CEMC exhibited a sharp decline in 2020 with the onset of the COVID-19 pandemic. In addition, analysis of relative proportions by anatomical region revealed that all five regions—ear, nose, pharynx, larynx, and oral cavity—similarly showed a precipitous decline in 2020. However, after 2021, the remaining four regions, excluding the larynx, demonstrated a gradual but incomplete recovery, plateauing at approximately 75% even into 2023 and 2024. In contrast, only the volume for laryngeal conditions showed a distinct recovery pattern, surpassing the pre-pandemic baseline level.

First, the sharp declines observed in 2020, as noted in many previous studies [1–22], can be attributed to multiple, interrelated factors. These include large-scale governmental infection-control measures, such as lockdowns and school closures; widespread adoption of personal preventive behaviors, including mask wearing and hand hygiene; a secondary reduction in the prevalence of upper respiratory tract infections resulting from these non-pharmaceutical interventions; and marked shifts in healthcare-seeking behaviors driven by public anxiety regarding COVID-19 transmission within medical environments.

In addition, the subsequent gradual recovery can be partially attributed to the progressive relaxation of the large-scale governmental infection-control measures and a concomitant reduction in public anxiety, which collectively improved medical access. Another principal factor is assumed to be an increase in the community-transmission of viral pathogens associated with upper airway infections, aligning with the normalization in social activities. However, shifts in healthcare-seeking patterns alone are considered unlikely to be the primary driver of this recovery, because emergency conditions generally manifest as intolerable symptoms that prevent patients from delaying medical intervention, regardless of the pandemic phase.

### 4.2 Variable Recovery in Infections Conditions by Anatomical region

Indeed, the present study demonstrated a similar trend in the relative proportions of acute infectious diseases when stratified by anatomical region, specifically demonstrating a sharp decline in 2020 and a subsequent incomplete return in the relative proportions of acute otitis media (AOM), acute sinusitis, and pharyngitis, and peritonsillar abscess. This persistent stagnation in the relative proportions of these acute infectious diseases even into 2024—a period by which large-scale infection-control measures had been substantially rescinded—is highly intriguing. This finding was ascribed to a continuous decrease in circulating respiratory pathogens in the surrounding urban community, driven by the sustained, habitual implementation of ethanol-based hand disinfection, frequent hand-washing, and face mask wearing among the general public.

In contrast, this study demonstrated a distinct trend in the relative proportion of laryngitis, which showed an outstanding surge surpassing the pre-pandemic baseline levels from 2022 onward. In addition, the relative proportion of epiglottic edema exhibited a transient surge only in 2021 followed by a return to the levels comparable with the remaining infectious diseases. These findings were ascribed to the direct impacts of COVID-19 infection. Especially, the post-pandemic surge in laryngitis surpassing the pre-pandemic baseline levels from 2022 onward can be plausibly related to the prominent laryngeal involvement associated with COVID-19. Previous reports have documented that SARS-CoV-2—particularly the Omicron variant, which became predominant from 2022 onward—can induce intense laryngeal inflammation, leading to prolonged dysphonia, laryngeal stenosis, and severe odynophagia [23–25]. Accordingly, the marked rebound in laryngitis after 2022 may reflect the widespread circulation of the Omicron variant in Osaka, as well as globally.

Similarly, the transient increase in epiglottic edema observed in 2021 may also have been associated with COVID-19–related upper airway inflammation [26,27]. However, in this study, SARS-CoV-2 antigen testing data could not be analyzed, as such information was unavailable in the medical records without explicit patient consent for research use.

### 4.3 Relationships of Fluctuations in Infectious Diseases to Adenotonsillar Hypertrophy

Our previous study [7] demonstrated that surgical referral volumes for adenoidal and tonsillar hypertrophy sharply declined immediately after the onset of the pandemic, remained suppressed during the mid-pandemic period, and subsequently showed an abrupt increase following the governmental decision to reclassify COVID-19 as a Category V infectious disease, with a sustained rise surpassing pre-pandemic levels. In addition, it was postulated that this mid-pandemic decline and post-pandemic surge in adenotonsillar hypertrophy were attributable not only to reduced healthcare utilization driven by infection-control measures and public anxiety regarding COVID-19 transmission, but also to a secondary reduction in pathogen exposure during the pandemic and a subsequent “catch-up” increase in circulating pathogens after public health restrictions were relaxed [7]. Given that lymphoid tissues respond dynamically to bacterial and viral stimulation, renewed exposure after prolonged suppression may induce sustained immunologic activation and promote lymphoid proliferation, thereby contributing to the development of adenotonsillar hypertrophy.

This high resolution and scale of this dataset derived from the CEMC provide the critical epidemiological and social missing link to answer the above core question raised in the introduction of the present study: how did upper respiratory tract infections that induce adenotonsillar hypertrophy actually behave in the community during and after the pandemic?

Critically, the present data demonstrate that upper airway infectious diseases (excluding laryngitis and epiglottic edema) declined sharply in 2020 and thereafter exhibited only gradual, incomplete recovery, plateauing at approximately 75 % of baselines without any “catch-up” overshoot, even in 2024 after the reclassification was implemented. This stands in marked contrast to the explosive post-restriction rebound observed in elective adenotonsillar surgery [7], suggesting that the emergency-care trajectory more faithfully reflects genuine pathogen dynamics, rather than patient hesitation to seek medical intervention.

### 4.4 Trends in Representative Otolaryngologic Emergency Conditions

Most representative emergency ENT diseases with the notable exception of SSNHL showed only a marginal decrease in 2020 and remained relatively stable thereafter. This divergence in trajectories between acute infectious diseases and these non-infectious emergency ENT conditions suggests that patients experiencing severe or intolerable symptoms—such as bleeding, disabling vertigo, or abnormal feelings due to foreign bodies—continued to seek emergency care at the CEMC even during the mid-pandemic period, despite concerns about COVID-19 exposure.

In contrast, SSNHL sharply declined to approximately 40% of the pre-pandemic baseline in 2020, followed by a persistent reduction through 2024. Several systematic reviews have summarized reported cases of SSNHL following COVID-19 infection [28–30]. However, a causal association between COVID-19 infection and SSNHL remains inconclusive, largely due to the limited number of cases and the predominance of case-based reports. Furthermore, SSNHL cases in the present study were not directly linked to confirmed COVID-19 infection, and notably, case volumes remained suppressed even in 2022, when the Omicron variant was widely circulating and COVID-19 incidence markedly increased nationwide. Accordingly, the post-pandemic reduction in the SSNHL volumes is highly unlikely to be related to COVID-19 infection dynamics.

SSNHL is a clinically diagnosed condition with a multifactorial and incompletely understood etiology, with proposed mechanisms including vascular compromise, autoimmune processes, viral infection, inner ear pathology, and central nervous system involvement [31,32]. However, definitive etiological factors are often difficult to identify in clinical practice, and the detection of SSNHL relies heavily on patient health-seeking behavior and timely access to otolaryngologic care. The sustained suppression of SSNHL may therefore reflect a composite of factors: possible genuine incidence decline, persistent alterations in care-seeking behavior, and disrupted referral pathways. Importantly, SSNHL—unlike epistaxis or pharyngeal foreign bodies—does not cause the kind of severe, urgent pain or bleeding that force patients to visit the emergency clinic immediately. Therefore, patients are much more likely to delay their hospital visits. The present aggregated dataset does not allow for the differentiation between true changes in disease incidence and shifts in healthcare utilization; thus, further studies incorporating individual-level clinical data will be required to clarify these mechanisms.

In addition, it is intriguing that epistaxis and pharyngeal foreign body showed an almost complete recovery to their baseline levels by 2023 thereafter. These results suggest that the incidence of epistaxis remains largely unaffected by daily lifestyle changes during the pandemic, regardless of whether it is idiopathic or hypertension-induced, and that accidental ingestion of fish bone—the most frequent causal agent for pharyngeal foreign bodies—gradually recovered, concurrently with the expanding opportunities gradually for dining out during the late phase of the pandemic.

### 4.5 Trends in Otolaryngologic Emergency Traumatic Conditions

Furthermore, new outpatients presenting with traumatic emergency conditions, stratified by otorhinolaryngologic region, showed a varied reduction by anatomical site, ranging from 53% to 81% of the baseline, followed by a gradual but incomplete recovery with variable fluctuations in subsequent years, reaching only 55% to 69% even in 2024. Because traumatic emergency conditions typically cause intolerable symptoms and injuries requiring urgent medical attention, most affected patients are unlikely to delay seeking care and are therefore expected to present promptly to the CEMC. Accordingly, the observed reduction in trauma-related case volumes following the onset of the pandemic likely reflects a genuine decrease in the incidence of traumatic events, potentially attributable to reduced social activity and mobility [33]. However, adequate quantitative evidence regarding social activity during the mid- and post-pandemic periods remains lacking. With regard to the causes of ENT traumatic conditions, the number of assault cases known to the Japanese police was reported to gradually decline by 2021, followed by a sharp and complete recovery in 2023 [34]. In contrast, the annual number of traffic accidents in Japan exhibited a marked decline during the early phase of the pandemic and was subsequently followed by a persistent plateau through 2023 [35]. Therefore, the trajectories of the volume of ENT traumatic conditions appear to differ by cause.

### 4.6 Limitations

This study has several limitations. A major limitation is that the CEMC is not a research-designated facility, and written informed consent for participation in medical research was not systematically obtained from patients. In particular, opt-out procedures—currently required even for retrospective observational studies in Japan—were not implemented. This institutional constraint precluded access to individual-level clinical data and restricted the analyses to aggregated, open-source statistics. Consequently, it was difficult to distinguish pediatric from adult patients in the emergency dataset. Although age-stratified analyses were not feasible from this dataset, clinical experience suggests that the majority of patients presenting to emergency services with AOM are likely to be children with severe otalgia or otorrhea, and that a substantial proportion of patients with acute rhinosinusitis or acute pharyngitis/tonsillitis are also pediatric. Accordingly, the observed trends in acute respiratory tract infections in the present study may partially reflect real-world changes in emergency otolaryngologic disease patterns across the pediatric population.

Second, as described above, clinicians at the CEMC are required to record only a single primary diagnosis for each visit, corresponding to the condition most strongly contributing to the patient’s presenting complaint. For example, in children presenting with AOM accompanied by nasal discharge, concomitant sinusitis is unlikely to be coded as the primary diagnosis. Consequently, this study carries an inherent bias where secondary diagnosis and comorbidities may be systematically underreported.

Third, acute conditions associated with severe or intolerable symptoms—such as AOM, pharyngitis, tonsillitis, peritonsillar abscess, laryngitis, and epiglottic edema—are inherently more likely to prompt urgent emergency visits. In contrast, patients with sinusitis lacking prominent facial pain often defer medical care until regular clinic hours. As a result, the number of sinusitis cases captured in the present dataset likely underestimates the true community burden of this condition. Consequently, the proportions of cases presenting to the CEMC may not uniformly reflect the underlying genuine prevalence of individual disease entities, but are rather skewed by the clinical severity of associated symptoms.

Fourth, it should be noted that the diagnostic category “catarrhal otitis media,” which corresponds clinically to otitis media with effusion (OME), was infrequently selected in the emergency setting. This terminology is historically outdated, rarely appearing in medical literature since the 1960s. Consequently, this under-selection is largely attributable to a profound lack of familiarity with this obsolete term, particularly among younger generations of attending physicians in routine emergency practice. Furthermore, OME is fundamentally not considered an emergency condition and rarely prompts urgent visits to emergency clinics owning to comparably tolerable symptoms such as hearing loss and ear fullness. Indeed, the annual number of cases recorded under the category “catarrhal otitis media” was remarkably small in the present dataset (fewer than 250 cases per year), standing in sharp contrast to acute otologic infections such as AOM, which ranged from 1,000 to 4,000 cases per year.

## 5. Conclusions

In conclusion, our analysis of volume changes in emergency otolaryngologic conditions at an urban emergency medical center addressed the primary question of whether acute respiratory tract infections surpassed pre-pandemic levels during the post-pandemic period. Nevertheless, our longitudinal findings demonstrated that acute respiratory tract infectious conditions experienced a sharper decline immediately after the onset of the pandemic than representative emergency otolaryngolgic conditions, including trauma. These findings likely reflect a genuine reduction in respiratory tract infection prevalence associated with public health measures. Although most infectious conditions (excluding laryngeal conditions) and traumatic conditions gradually but incompletely recovered to near pre-pandemic baseline levels during the mid- and post-pandemic periods, laryngeal conditions exhibited a unique post-pandemic increase, potentially related to COVID-19 infection. In contrast, most other representative emergency otolaryngologic conditions remained relatively stable throughout the mid- and post-pandemic phases; especially, epistaxis and pharyngeal foreign bodies showed an almost complete recovery. However, SSNHL remained persistently reduced for reasons that remain to be fully elucidated. The present study provides valuable real-world evidence to inform future health-care planning in urban otolaryngologic emergency settings and offers a robust framework for understanding how large-scale pandemic and subsequent public health crises can differentially disrupt emergency otolaryngologic care.

## Data Availability

All data produced in the present study are available upon reasonable request to the authors

## Acknowledgements

The author expressed gratitude to the Osaka Association of Otorhinolaryngologists for their generous support and cooperation in data collection for this study.

## Ethical issues

The study protocol complied with the Declaration of Helsinki, and was approved by the Institutional Review Board of Osaka City General Hospital (approval number: 2405013).

## Written consent for publication

Not applicable. This study exclusively utilized publicly available, aggregated open-source data from annual statistical reports, and no individual-level identifiable patient data were included.

## Declaration of generative AI and AI-assisted technologies in the writing process

During the preparation of this work, the author (MO) utilized generative AI tools, including ChatGPT-5.5, Gemini 2.0 and Claude Sonnet 4.6. These tools were used strictly to assist with English language polishing for enhancing the readability and evaluate alternative phrasing for clinical descriptions. Following the automated suggestions, the author thoroughly reviewed, selected, and edited the text to ensure precise logical alignment with the study results. The author retains full responsibility for the final content of the published article.

## Author contributions

MO conceptualized, organized and coordinated the present study and contributed to data curation, visualization, and manuscript writing.

